# Novel ELISA protocol links pre-existing SARS-CoV-2 reactive antibodies with endemic coronavirus immunity and age and reveals improved serologic identification of acute COVID-19 via multi-parameter detection

**DOI:** 10.1101/2020.09.15.20192765

**Authors:** Rachel R. Yuen, Dylan Steiner, Erika L. Smith, Riley M.F. Pihl, Elizabeth Chavez, Alex Olson, Lillia A. Baird, Filiz Korkmaz, Patricia Urick, Manish Sagar, Jacob L. Berrigan, Suryaram Gummuluru, Ronald B. Corley, Karen Quillen, Anna C. Belkina, Gustavo Mostoslavsky, Wenda Gao, Ian Rifkin, Amedeo J. Cappione, Nina H. Lin, Yachana Kataria, Nahid Bhadelia, Jennifer E. Snyder-Cappione

## Abstract

The COVID-19 pandemic has drastically impacted work, economy, and way of life worldwide in 2020. Sensitive measurement of SARS-CoV-2 specific antibodies would provide new insight into pre-existing immunity, virus transmission dynamics, and the nuances of SARS-CoV-2 pathogenesis. To date, existing SARS-CoV-2 serology tests have limited utility due to insufficient reliable detection of antibody levels lower than what is typically present after several days of symptoms. To measure lower quantities of SARS-CoV-2 IgM, IgG, and IgA with higher resolution than existing assays, we developed a new ELISA protocol with a distinct plate washing procedure and timed plate development via use of a standard curve. This ‘BU ELISA’ method exhibits very low signal from samples added to buffer coated wells at as low as a 1:5 dilution. Use of this method revealed circulating SARS-CoV-2 receptor binding domain (RBD) and nucleocapsid protein (N) reactive antibodies (IgG, IgM, and/or IgA) in 44 and 100 percent of pre-pandemic subjects, respectively, and the magnitude of these antibodies tracked with antibody levels of analogous viral proteins from the 229E and/or NL63 endemic coronavirus (eCoV) strains. The disease status (HIV, SLE) of unexposed subjects was not linked with SARS-CoV-2 reactive antibody levels; however, quantities were significantly lower in subjects over 70 years of age compared with younger counterparts. Also, we measured SARS-CoV-2 RBD- and N-specific IgM, IgG, and IgA antibodies from 29 SARS-CoV-2 infected individuals at varying disease states, including 10 acute COVID-19 hospitalized subjects with negative serology results by the EUA approved Abbott IgG chemiluminescent microparticle immunoassay. Measurements of SARS-CoV-2 RBD- and N-specific IgM, IgG, IgA levels measured by the BU ELISA revealed higher signal from 9 of the 10 Abbott test negative COVID-19 subjects than all pre-pandemic samples for at least one antibody specificity/isotype, implicating improved serologic identification of SARS-CoV-2 infection via multi-parameter, high sensitive antibody detection. We propose that this improved ELISA protocol, which is straightforward to perform, low cost, and uses readily available commercial reagents, is a useful tool to elucidate new information about SARS-CoV-2 infection and immunity and has promising implications for improved detection of all analytes measurable by this platform.

## Introduction

From the first reported case of COVID-19 caused by the virus SARS-CoV-2 in December 2019 ^1,2^ there have been more than 76 million reported cases and 1.69 million deaths worldwide as of December 20, 2020. Common symptoms of SARS-CoV-2 infection include fever, cough, myalgia, and fatigue and these symptoms vary widely in magnitude, nature, and duration between individuals for reasons that are not clear to date ^3,4^, with some individuals with confirmed infections remaining asymptomatic ^5^. Epidemiological evidence indicates silent viral spread via asymptomatic individuals within communities and the extent of this form of transmission is currently unclear ^6^. SARS-CoV-2 has homology to other alpha and beta ‘common cold’ endemic coronaviruses (eCoVs) in circulation, and cross-reactive T cell immunity to SARS-CoV-2 spike (S) and nucleocapsid (N) proteins are present in a substantial percentage of unexposed individuals ^7-10^. Also, reactive antibodies to SARS-CoV-2 S and N proteins are present in unexposed individuals, with virus neutralization activity reported from pre-pandemic pediatric samples ^11,12^. It is postulated that this cross-reactive immunity may influence the nature and severity of COVID-19 symptoms upon infection and impact disease course ^13^ and may impact herd immunity.

Sensitive and accurate detection of virus-specific immune factors, such as antibodies, is imperative in order to measure rates of SARS-CoV-2 infections within communities with greater accuracy, to more fully define cross-reactive immunity in unexposed individuals, and to gain new understanding about the nature of effective versus potentially deleterious immune responses upon SARS-CoV-2 exposure. Antibody measurements are of particular importance, as pathogen-specific immunoglobulins are a known first line of defense upon exposure and can prevent new infections. Antibody titers are used to assess both likelihood of protection from re-infection and general vaccine efficacy ^14^. A variety of SARS-CoV-2 serological assays have been developed by multiple manufacturers and academic institutes and many are CE-marked and granted emergency use authorization (EUA) from the US Food and Drug Administration (FDA). Varieties include point-of-care rapid lateral flow assays, chemiluminescence immunoassays (CLIA), multi-plex bead/cell based-assays^15,16^, and enzyme-linked immunosorbent assays (ELISA)^17-20^. These tests detect antibodies that primarily target the nucleocapsid protein (N) or the spike (S) protein of SARS-CoV-2, and specifically the Receptor Binding Domain (RBD) of spike which is an immunodominant surface protein targeted by neutralizing antibodies and a main target antigen for vaccine development ^20-22^. Some of these tests possess high sensitivity and specificity for detection of SARS-CoV-2 antibodies 14 days after diagnosis and/or symptom onset ^23-26^. However, others report negative results from individuals who are asymptomatic, mildly symptomatic, or symptomatic for less than 14 days, even when SARS-CoV-2 infection is confirmed ^19,27,28^; whether such individuals possess antibodies below the limit of the detection of the particular test used or lack these antibodies altogether is unresolved.

To enable detection of low levels of SARS-CoV-2-reactive antibodies, we modified the standard ELISA procedure, particularly the plate washing method, to improve sensitivity. Our protocol (the ‘BU ELISA’) allows clear SARS-CoV-2-reactive antibody signal resolution at sample dilutions as low as 1:5. Using this protocol we measured the levels of SARS-CoV-2-reactive IgM, IgG, and IgA from plasma or serum from three groups of individuals: (1) 71 subjects that varied by age, HIV infection, and systemic lupus erythematosus (SLE) disease status with all samples collected before November 8^th^, 2019 (‘pre-pandemic’); (2) 20 subjects hospitalized with COVID-19 (‘Acute’) (3) nine subjects with samples collected three-six months after confirmed SARS-CoV-2 infection (‘Convalescent’). In addition, the performance of the BU ELISA, Antagen’s IgM IgG Lateral Flow Device (LFD) test and the Abbott IgG chemiluminescent microparticle immunoassay (CMIA) were directly compared from samples from the three subject groups.

## Results

### A modified ELISA protocol demonstrates low noise from high concentration human serum and plasma samples

The Enzyme-Linked Immunosorbent Assay (ELISA) is a commonly used method for the measurement of analytes in a suspension sample. While low cost and easy to adapt in most lab settings, a limitation of this platform is high background from some biological samples at low sample dilutions. Specifically, optical densities (ODs) from sample dilutions lower than 1:100 is often sizeable and can mask the analyte of interest. This issue is particularly germane to serologic testing for SARS-CoV-2, as antibodies that are cross-reactive in unexposed individuals, newly generated in asymptomatic and/or recent infections, induced from an encounter with low viral dose, or waned post convalescence may be missed because levels are below the limit of detection of current assays. To address this issue, we have developed an ELISA protocol with unique steps to reduce non-specific signal at low sample dilutions. One change is the plate washing procedure, which is performed manually by an operator using a multichannel pipettor and includes agitation and soaking steps with repeated complete removal of residual fluid as described (Methods and Supplemental Figure 1). ELISAs were performed that compared buffer coated well OD values of five human plasma samples with plates washed with our method or an automated plate washer and the total levels of non-specifically bound IgG was determined. The manual washing procedure resulted in a notably lower average and range of ODs at 1:5, 1:10, and 1:25 dilutions as compared with the automated washer (Figure 1A). This BU ELISA protocol was run on samples from a total of 71 pre-pandemic and 29 SARS-CoV-2-infected subjects (Table 1), with paired antigen coated and buffer coated wells for six or seven sample dilutions (Supplemental Figure 1) for all subjects. The average ODs for buffer coated, 1:5 diluted sample loaded wells from all subject samples measured at this dilution were 0.16, 0.098, and 0.076 for IgM, IgG, and IgA respectively (Figure 1B). Given these low background OD values and the results from the wash method comparison, it’s possible that details of our protocol other than the washing method may contribute to these low background ODs, such as the type of plates, the blocking buffer/sample diluent used, and the number and placement of washing steps (Methods and Supplemental Figure 1). This buffer only coat ‘noise’ is remarkably consistent between multiple runs of a given sample (Supplemental Figure 2) and appears to be due to components within the sample, such as IgG and inflammatory factors^29^ and not due to assay variability. Importantly, when ODs from uncoated wells with the same dilution of sample are not measured and properly subtracted, incorrect interpretation of results as positive can occur^30^; therefore, the no coat values were subtracted from coated OD results for all results to determine the true antigen-specific signal. Also, detection antibodies were tested for specificity to confirm accuracy of isotype-specific readouts and the ability of our IgG detection reagent to measure all four IgG subclasses (Supplemental Figure 3).

**Table 1.** Cohort characteristics

| Cohort Characteristics | Age (average, range) | Sex, M (%) | Length of Symptoms<br>(days average, range) |
| --- | --- | --- | --- |
| **Pre-pandemics** (n = 71) |  |  |  |
| Healthy (n = 37) | 50 (21 - 96) | 78 | N/A |
| HIV+ (n = 24) | 45 (22 - 79) | 96 | N/A |
| SLE (n = 10) | 39 (23 - 69) | 30 | N/A |
| **Acute** (n = 20) | 63 (48 - 84) | 80 | 18 (3 - 40)<sup>+</sup> |
| **Convalescent** (n = 9) | 53 (35 - 77) | 22 | 27 (0 - 61)* |

+ current length of symptoms at time of sample collection
\* total length of symptoms

**Figure 1.**
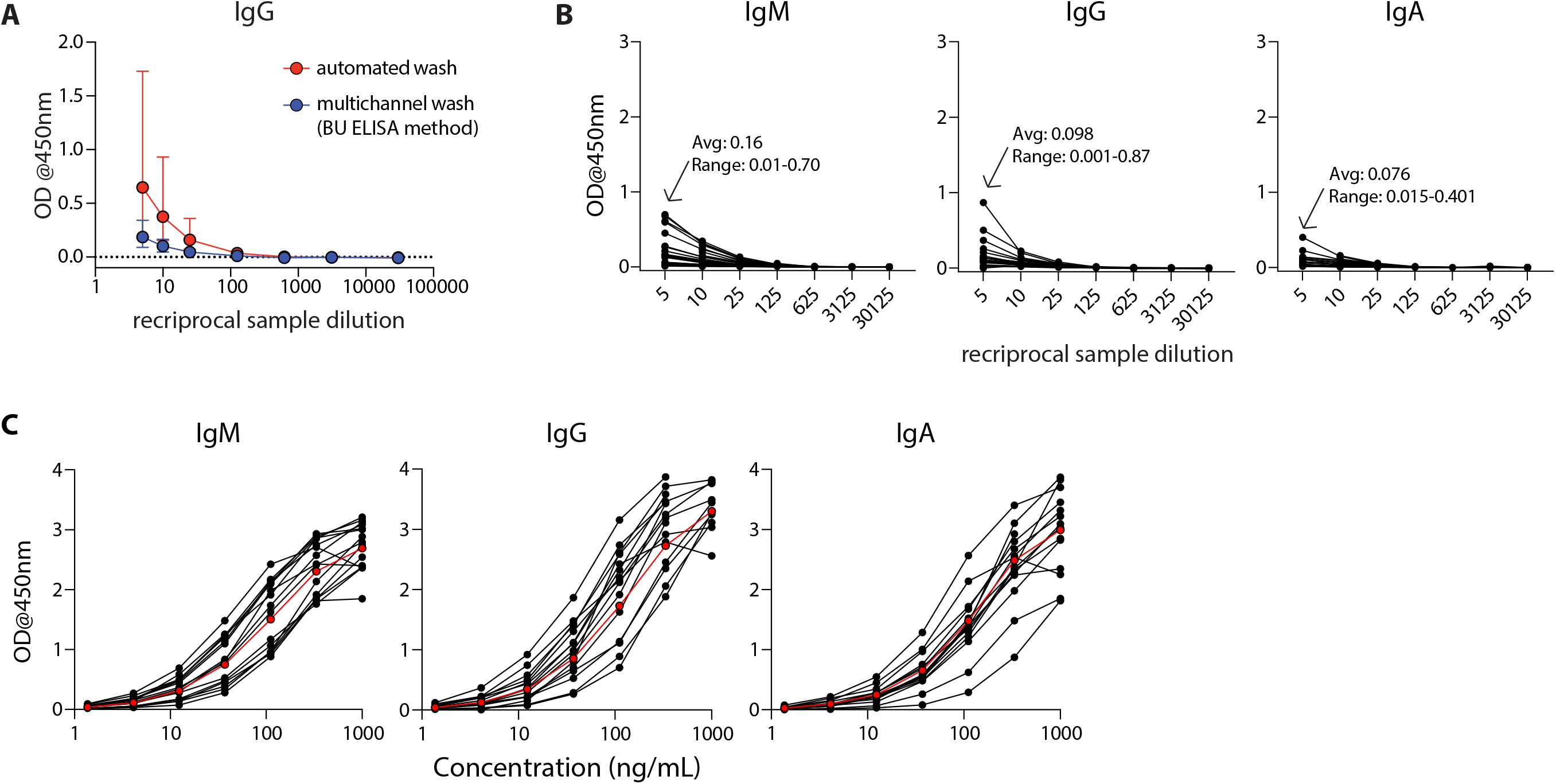
The modified ELISA (BU ELISA) protocol exhibits low background signal at high sample concentration and use of SARS-Cov-2 RBD-recombinant antibody standard curves allows for accurate sample quantification via accounting for OD drift between experimental runs. (A) Dilution curves of buffer only coated wells from five donor samples after using an automated plate washer or the BU ELISA method of multichannel plate washing. Experiment was performed once. (B) Representative dilution curves of buffer only coated wells from 30 subjects, average and range of 1:5 sample dilution for each isotype from all subjects; IgM, IgG, and IgA were detected in individual assays. (C) Representative IgM, IgG, and IgA standard curves from 15 different experimental runs are shown. The average of all runs shown in red.

**Figure 2.**
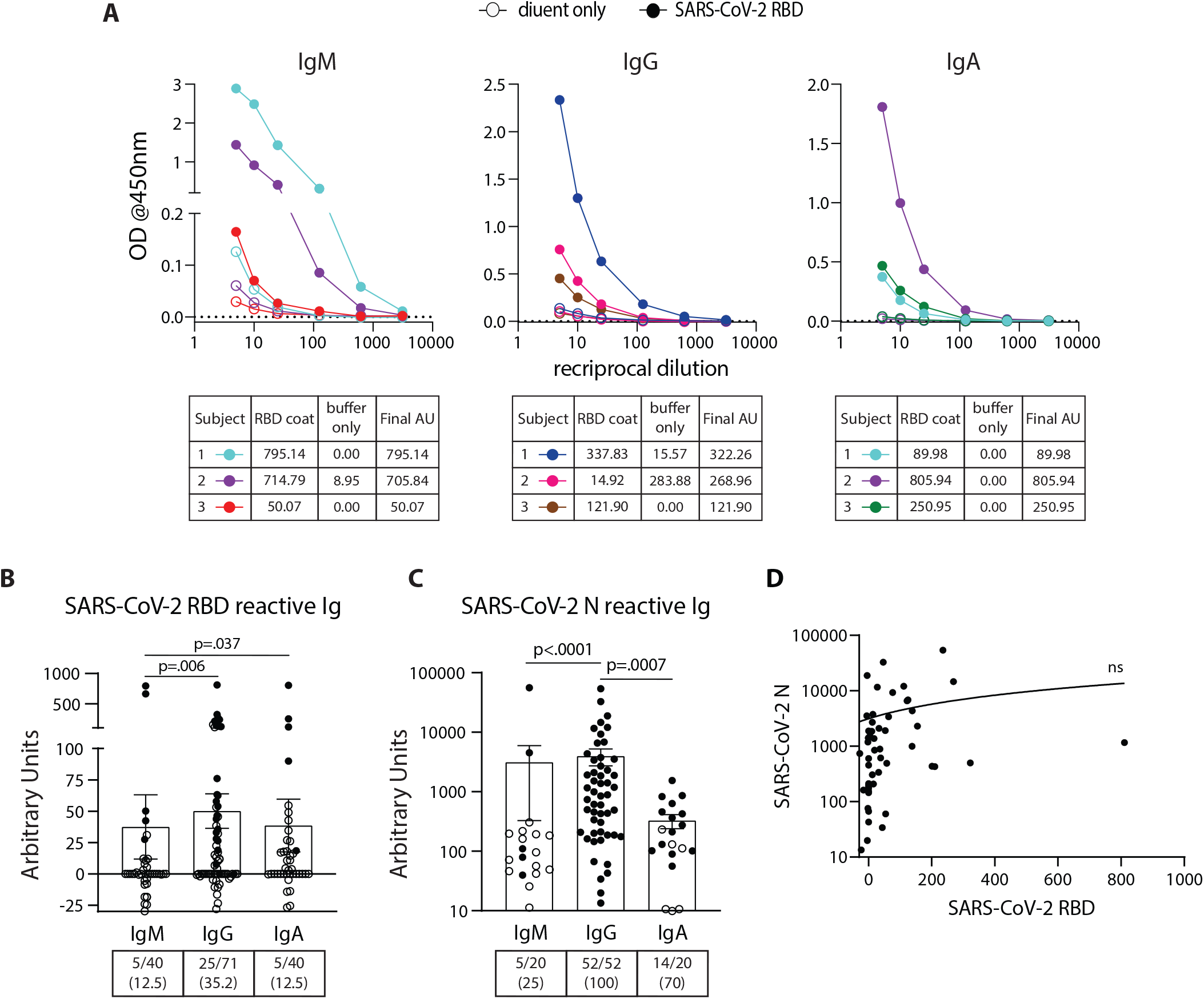
Detection and quantification of SARS-CoV-2 RBD- and N-reactive antibodies in pre-pandemic samples. (A) Representative dilution curves of three pre-pandemic samples for each isotype for SARS-CoV-2 RBD reactive Ig. Open and solid symbols represent buffer only coat and SARS-CoV-2 RBD coat, respectively. Arbitrary Units (AU) were calculated as described in Methods and shown beneath the respective isotype graph for buffer only and SARS-CoV-2 RBD coat. AUs for SARS-CoV-2 RBD (B) and N (C) reactive IgM, IgG, and IgA in pre-pandemic samples. Open and solid symbols represent negative and positive results, respectively, as determined by Metric 1. Enumeration of the positive samples for each isotype in the pre-pandemic cohort is shown beneath each graph with percentages of total in parentheses. (D) Correlation between AUs for IgG reactive to SARS-CoV-2 RBD and N (n = 53). Statistical analyses were performed using an unpaired non-parametric Mann-Whitney t-test and Pearson’s rank test.

**Figure 3.**
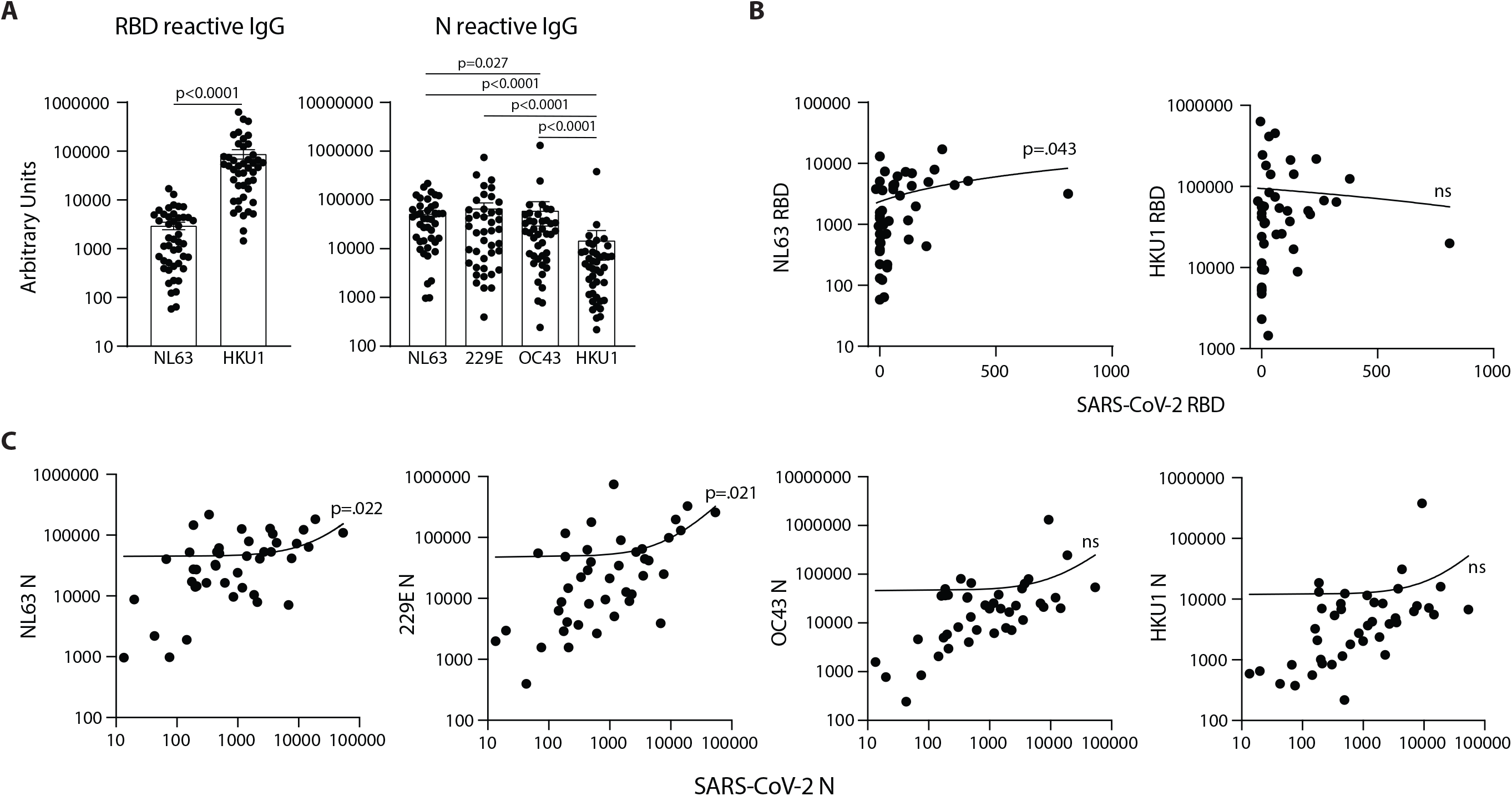
SARS-CoV-2 RBD and N reactive IgG in pre-pandemic samples track with IgG recognizing analogous proteins of the eCoV strains NL63 and 229E. (A) AUs of IgG reactive to RBD of NL63 and HKU1 and N of all four eCoV strains (NL63, 2293, OC43, and HKU1). (B) Correlation between SARS-CoV-2 RBD IgG levels with NL63, HKU1 RBD IgG levels in individual subjects. (C) Correlation between SARS-CoV-2 N IgG and NL63, 229E, OC43, and HKU1 N IgG levels, n = 42-45. Statistical analyses were performed using an unpaired non-parametric Mann-Whitney t-test and Pearson’s rank test in GraphPad Prism.

### Modification of ELISA development duration based on standard curve signal detection enables accurate comparison of antibody levels between experimental runs by minimizing impact of OD drift

During assay development we noted differences in OD values in different experimental runs even with strict adherence to all procedures and length of steps. Therefore, for all sample runs, we included a standard curve using recombinant monoclonal IgM, IgG, and IgA antibodies that recognize SARS-CoV-2 RBD for each of the respective isotype assays and stopped the development reaction when there was a visible difference between the seventh dilution (1.37ng/ml) of the standard and the ‘zero’ (sample diluent only) well. Addition of these standards and timing of development in this manner helped to ensure accurate calculation of the relative antibody levels (termed ‘Arbitrary Units’ on a ng/ml scale, calculated as described in Methods) between samples run on different days, plates, and/or by different operators. The OD values of the standard curves following this development procedure for the IgM, IgG, and IgA assays for 15 representative runs are shown (Figure 1C). The development time of these runs to complete visualization of the standard curve development ranged from ∼8-30 minutes, demonstrating the need to adjust substrate incubation time per experimental run to maximize signal detection.

### SARS-CoV-2 RBD-reactive antibodies were detected at low levels in 44 percent of pre-pandemic samples

SARS-CoV-2 RBD IgM, IgG, and IgA ELISA assays were performed on 40, 71, and 40 pre-pandemic samples, respectively (Table 1) using the BU ELISA protocol. The OD curves from the BU ELISA for both the buffer coat and SARS-CoV-2 RBD coated wells (after first subtracting the blank well(s) with paired coat) from seven pre-pandemic subjects for IgM, IgG, and/or IgA is shown (Figure 2A). There is clear RBD-specific signal with with a linear loss of OD with sample dilution, providing evidence of true specific signal (Figure 2A). The calculated Arbitrary Units (AUs) from the buffer only and antigen coated wells from these curves is shown beneath each respective isotype graph. We defined a subject as positive for a given antibody readout as follows: the OD value from the RBD-coated well ≥2.5x the uncoated well from the paired sample dilution for at least two dilutions in the series and ≥0.1 for at least one dilution. Following this guideline, 31/71 of the unexposed individuals possessed reactive antibodies of at least one isotype to SARS-CoV-2 RBD, albeit all at very low levels in the circulation (∼40 ng/ml) (Figure 2B). We compared the calculated AUs from IgG reactive to SARS-CoV-2 Spike (S) and RBD from 14 pre-pandemic subjects and found no significant correlation (Supplemental Figure 4). These results could be due to differences in portions and/or presentation of the RBD antigen in the different tests.

**Figure 4.**
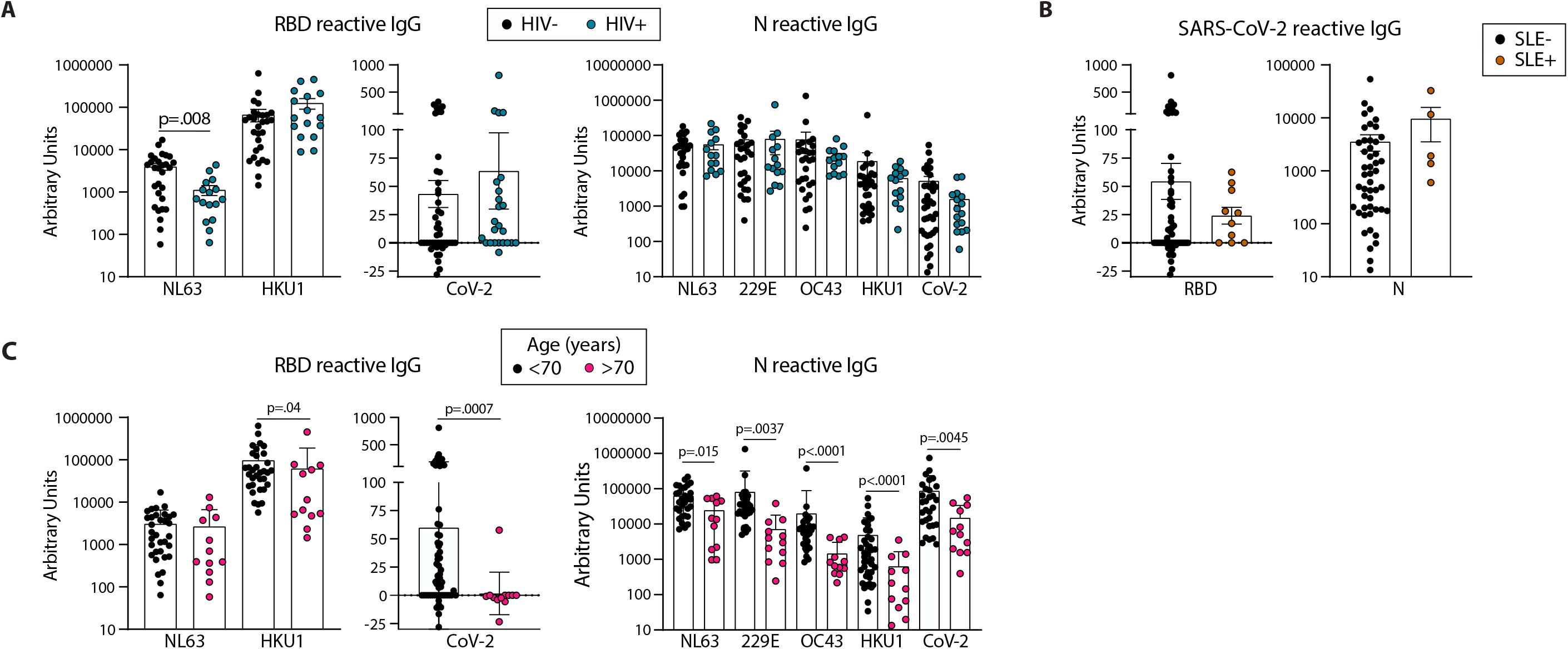
Older age is associated with lower circulating antibodies reactive with SARS-CoV-2 and eCoV RBD and N antigens. Quantification of IgG reactive to RBD of NL63, HKU1, and SARS-CoV-2 and N of NL63, 229E, OC43, HKU1, and CoV-2 in pre-pandemic samples regrouped based on HIV (A) or SLE (B) disease status or age (C). Statistical analyses were performed using an unpaired non-parametric Mann-Whitney t-test.

### All pre-pandemic subjects contained circulating SARS-CoV-2 nucleocapsid (N) IgG and/or IgA antibodies with a wide range of levels found between individuals

Using the BU ELISA protocol, we measured IgM, IgG, and IgA levels reactive with SARS-CoV-2 nucleocapsid protein (N) from 20, 53, and 20 subjects from our pre-pandemic cohort, respectively. Seven dilutions were run for all samples with and without N coated wells as with the SARS-CoV-2 RBD assays; sample dilution curves were generated, and positive results were determined using Metric 1 and AUs calculated. A wide range of pre-existing antibody levels were found; for example, the IgG levels range from 0.0134 to 54 μg/ml (Figure 2C).

### No correlation between levels of cross-reactive SARS-COV-2 antibodies to RBD and N antigens

We compared the levels of SARS-COV-2 RBD- and N-reactive antibodies between individual pre-pandemic subjects in our cohort and found no correlation between the two readouts (Figure 2D). This suggests these antibodies to different portions of SARS-CoV-2 are elicited during distinct immune responses.

### IgG reactive with SARS-CoV-2 RBD and N in pre-pandemic samples correlate with immunity to NL63 and both NL63 and 229E eCoV strains, respectively

To determine if antibodies elicited by endemic coronavirus (eCoV) infections are linked to antibodies reactive to SARS-CoV-2 in unexposed subjects, IgG specific for NL63 and HKU1 RBD proteins, and IgG reactive with the N protein from all four eCoV strains in the circulation (NL63, HKU1, 229E and OC43) were measured. The levels of antibodies to eCoV RBD and N proteins in pre-pandemic samples showed general differences, with more IgG reactive to HKU1 than NL63 RBDs among the subjects and similar levels of IgG reactive with N proteins of the NL63, 229E, and OC43 strains, with lower levels reactive with the HKU1 N (Figure 3A). IgG reactive with SARS-CoV-2 RBD significantly correlated with NL63 and not HKU1 RBD-specific IgG (Figure 3B) and IgG reactive with SARS-CoV-2 N correlated with NL63 and 229E but not HKU1 or OC43 N-specific IgG (Figure 3C). Taken together, these results suggest previous coronavirus infections with the NL63 and 229E strains elicit SARS-CoV-2 cross-reactive antibody immunity.

### HIV or SLE disease status does not impact SARS-CoV-2 reactive RBD and N antibody levels in unexposed individuals

We next compared the levels of eCoV and/or SARS-CoV-2 reactive antibodies in our pre-pandemic cohort with the subjects re-classified by HIV and SLE status. We found lower levels of NL63 RBD-reactive IgG in HIV+ as compared to uninfected subjects (Figure 4A); however, there were no other differences found between the antibody levels reactive to the RBD or N proteins, for either the eCoV strains or SARS-CoV-2, between groups classified via HIV or SLE status (Figure 4A, B).

### Unexposed individuals over 70 years old have significantly lower levels of SARS-CoV-2 RBD and N reactive IgG than younger counterparts

We next re-categorized our pre-pandemic cohort into two groups by age, <70yo (n=29-59) and >70 yo (n=12). All eCoV and SARS-CoV-2 reactive IgG levels measured were lower in the >70yo group, with high significance for SARS-CoV-2 RBD (p=.0007) and N (p=.0045). It should be noted that comparisons of younger (<35yo) and middle aged (40-65yo) groups did not yield notable differences (data not shown). Taken together, these results suggest that age may impact the magnitude of eCoV and SARS-CoV-2 cross-reactive antibody immunity more than a chronic viral infection (HIV) or an autoimmune disease (SLE).

### Comparison of SARS-COV-2 specific RBD and N antibody levels between hospitalized COVID-19 subjects with acute disease and convalescent survivors of infection

We next used the BU ELISA to measure the IgM, IgG, and IgA RBD- and N-specific antibodies from individuals at different times and magnitudes of severity after SARS-CoV-2 infection. Of the 20 COVID-19 hospitalized subjects (Acutes), 10 scored negative and 10 positive on the EUA approved Abbott SARS-CoV-2 N-specific IgG CMIA. RBD- and N-specific IgM and IgA was higher in all Acutes as compared with the Convalescent subjects, but IgG levels were similar, suggesting waning of IgM and IgA over time or reduced induction of these isotypes in subjects that do not require hospitalization (Figure 5A). Also, RBD- and N-specific IgG levels significantly correlate among all COVID-19 subjects in the study (n=29) and IgM and IgA also trend in a similar manner (Figure 5B).

**Figure 5.**
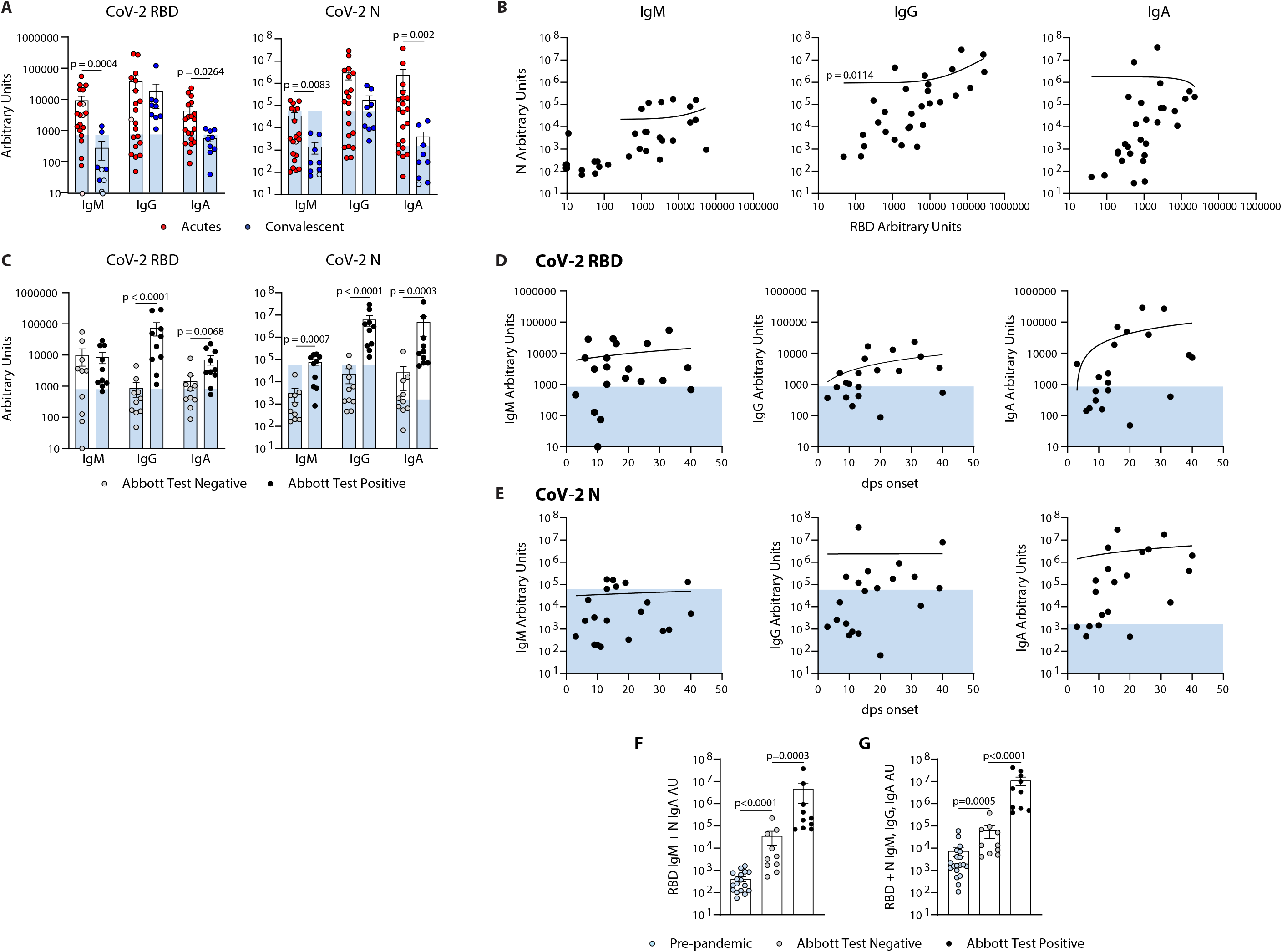
Quantification of the relative levels of IgM, IgG, and IgA-reactive SARS-CoV-2-RBD and N antibodies from acute and convalescent SARS-CoV-2 infected subjects. (A) Arbitrary Units (AUs) of SARS-CoV-2 RBD and N reactive IgM, IgG, and IgA of acute and convalescent subjects. Open and solid symbols represent negative and positive results, respectively, as determined by our Metric 1 described in Methods. (B) Correlation between SARS-CoV-2 RBD and N IgM, IgG, and IgA AUs. (C) Quantification of SARS-CoV-2 RBD and N reactive IgM, IgG, and IgA of acute subjects regrouped based on results from Abbott’s SARS-CoV-2 IgG CMIA. Correlation between SARS-CoV-2 RBD (D) and N (E) IgM, IgG, and IgA AUs with the number of days post symptom (dps) onset at time of sample collection for acute subjects. Quantification of SARS-CoV-2 RBD reactive IgM and N reactive IgA (F) and RBD & N reactive for IgM, IgG, and IgA (G) for pre-pandemics (n = 19) and Acutes re-classified based on Abbott test results. Light blue bars depict AU range of pre-pandemics for each respective antigen and isotype. Statistical analyses were performed using an unpaired non-parametric Mann-Whitney t-test and Pearson’s rank test.

### Comparison of RBD- and N-specific antibody levels measured by the BU ELISA from hospitalized, acute COVID-19 subjects with positive versus negative Abbott test results

Next, we compared the RBD- and N-SARS-CoV-2 specific antibody levels detected from the BU ELISA between Acute subjects with negative or positive Abbott IgG test results. The BU ELISA detected reactive antibodies from all samples, with all the Abbot test positive subjects with AU values above the pre-pandemic range for RBD and N-specific IgG (Figure 5C). The AU values for six Abbott test negative subjects were above the pre-pandemic range for RBD-specific IgM and for N-specific IgA, and two Abbott test negative subjects have RBD-specific IgG above the pre-pandemic range (Figure 5C), indicating serological evidence of infection in many of these subjects.

### Evidence of diversity of adaptive B cell response induction among acutely infected, hospitalized COVID-19+ subjects

Next, we compared the levels of RBD- and N-reactive IgM, IgG and IgA from our cohort of acute COVID-19 subjects, and found general trends showing higher antibody levels with more days of symptoms; however, some subjects have pre-pandemic levels of RBD and/or N-reactive IgG, even after as long as 40 days symptomatic (Figure 5D, E).

### Combinational analysis of readouts by the BU ELISA reveal SARS-CoV-2 reactive antibody levels are significantly higher in acutely infected COVID-19+ subjects with negative Abbott test results than pre-pandemics

Next, we compared the combined AU values of both RBD-reactive IgM and N-reactive IgA and all six readouts performed and found significant differences between the pre-pandemic and acute Abbott test negative groups, as well as between acute Abbott test negative versus positive groups (Figures 5F, G). These results indicate that multi-parameter detection, comprised of multiple isotypes and antigen reactivities from a sensitive serology test, could improve serologic diagnostics of SARS-CoV-2 infection.

### Three-way comparison of the BU ELISA results with the Antagen LFD and Abbott CMIA reveal 9 of 10 Abbott test negative COVID-19 subjects exhibit SARS-CoV-2 specific antibody levels above all pre-pandemic samples for at least one readout by the BU ELISA

The AU values from 10 pre-pandemic subjects with highest AU values for SARS-CoV-2 RBD and/or N-reactive IgG were directly compared with results from the Abbott CMIA assay and a lateral flow rapid test (Antagen Pharmaceuticals). The Abbott CMIA test measures IgG reactive to SARS-CoV-2 N, and the Antagen LFD test measures SARS-CoV-2 RBD-reactive IgG and IgM. Both commercial tests detected no SARS-CoV-2 reactive IgG in the pre-pandemic samples, and correctly identified the infection status of all subjects within the convalescent group (Table 2). Among the Acute subjects, of the 10 samples that scored positive by the Abbott test, they were also positive by the LFD test for IgG. Of the 10 Acute subjects that scored negative on the Abbott test, the LFD test successfully identified 5/10 subjects as SARS-COV-2 antibody positive for IgG and/or IgM (Table 2). Also, nine of 10 acute COVID-19 subjects that scored negative on the Abbott test have AU values above all pre-pandemics tested for at least one of the six BU ELISA readouts (all but Subject A4).

**Table 2.**
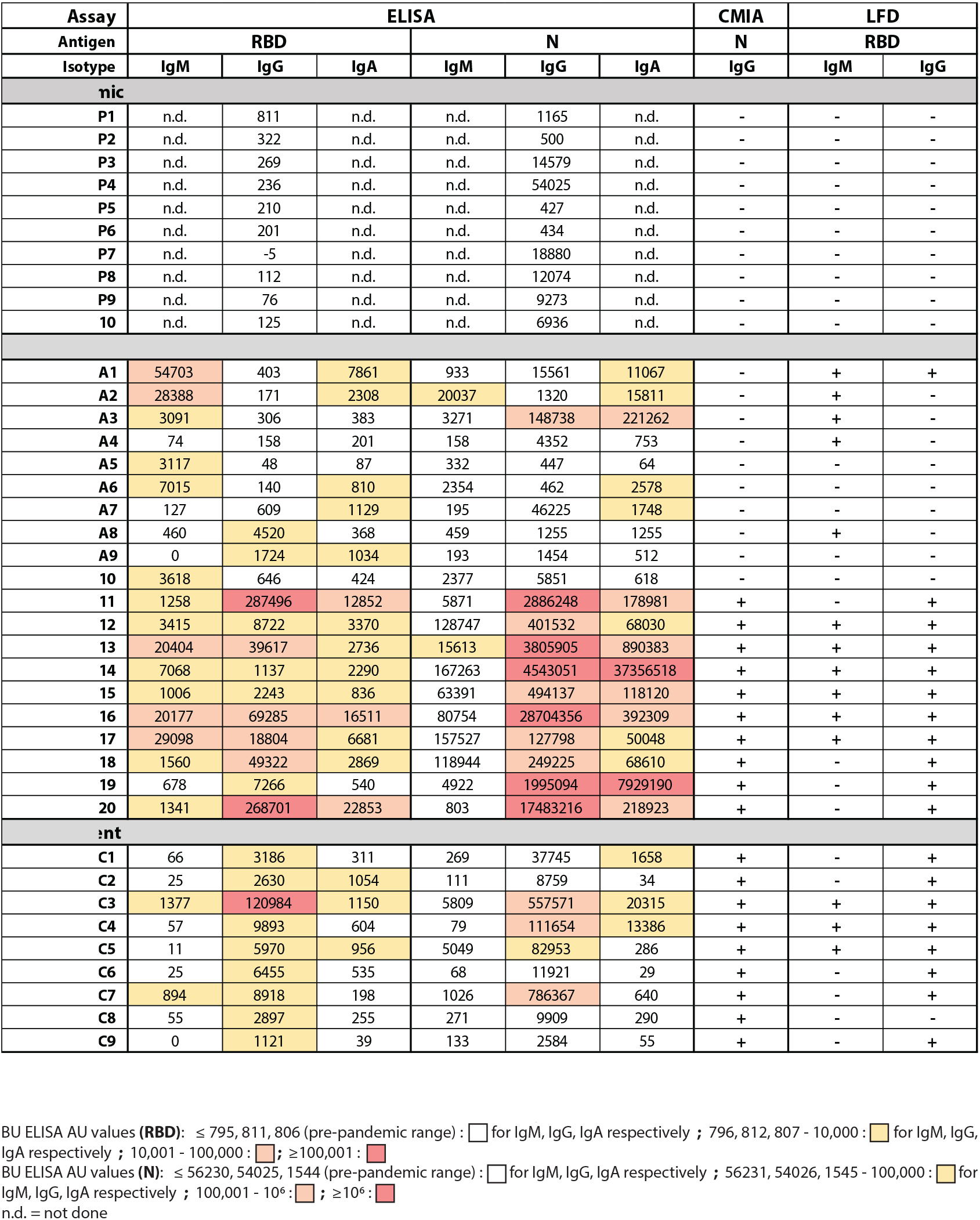
Comparison of SARS-CoV-2 reactive antibody results measured by the BU ELISA protocol, the EUA approved Abbott IgG chemiluminescent microparticle assay, and Antagen’s lateral flow rapid test.

## Discussion

Accurate and sensitive measurement of virus-specific antibodies could complement diagnostic testing, provide information about the true prevalence of infection, provide insight into anti-viral immunity, and help assess vaccine responses. However, a lack of required sensitivity and specificity of many of the SARS-CoV-2 antibody tests available to date have led some to conclude that they have limited clinical utility in combating COVID-19 ^31^. Here, we present a modified ELISA protocol with exceptional sensitivity with high concentration samples that enables the detection of low levels of antigen-specific antibodies in human specimens.

The BU ELISA is straightforward, comprised of reagents that are readily available from commercial vendors and can easily be adapted for other applications and analytes. However, a limitation of this assay is that it is currently considerably lower in throughput compared to other serological platforms. This protocol requires an operator for the manual wash steps, limiting the number of plates that can be run compared to automated methods, however, there is potential for throughput increase if automated washers/ELISA systems can be adapted to more closely mimic this protocol.

Other important features of our approach include the inclusion of paired sample dilutions with buffer only coated wells to enable detection of true antigen-reactive signal and adjustment of the length of substrate incubation time based on standard curve development for OD standardization to enable direct comparison between samples on different plates. Quantification of relative antibody levels via Arbitrary Units (AU) or a similar method will be imperative for determining which convalescent samples have antibody levels sufficient for effective plasma transfer as well as other applications. However, while we believe this is a preferred approach for determining of relative output values within all samples, it is critical to note that the unique dynamics of the panoply of antibodies of varying affinities and isotypes within a given specimen causes inherent confounding factors to serologic readouts. For example, a specimen with a high level of SARS-CoV-2 RBD reactive IgM antibodies could have a lower detected signal for IgG and IgA due to IgM’s pentameric conformation blocking many binding sites. Also, higher affinity antibody clones (IgG and IgA vs IgM, for example) may outcompete for binding sites of the coated antigen and thereby be detected more readily than others. We can account for this issue to some extent via measurement of all three major isotypes in all samples.

Here, we report links between antibody responses to endemic coronavirus (eCoV) strains and levels of cross-reactive antibodies with SARS-CoV-2 in unexposed individuals. These results support recent reports of SARS-CoV-2 T cell responses correlating with eCoV T memory in pre-pandemic samples.. Importantly, hospitalized COVID-19 subjects with recent eCoV infections were significantly less likely to require the ICU or die ^32^, and blood samples from virally unexposed children were found to possess neutralization activity against SARS-CoV-2 ^11^. These results collectively implicate past eCoV infections with protection from severe outcomes to COVID-19 due to cross-reactive immunity, including antibodies. We found widely varying quantities of SARS-CoV-2 reactive N IgG within our pre-pandemic cohort, an agreement with another study ^12^ and reports of N-specific T cell immunity in unexposed subjects ^7-10^. The magnitude and signature of one’s eCoV immunity may partially explain the profound diversity of outcomes that occurs upon SARS-COV-2 infection. Therefore, screening for this cross-reactive immunity may provide new insight into an individual’s risk of serious COVID-19.

44 percent of the pre-pandemic subjects possessed antibodies that bind to SARS-CoV-2 RBD, albeit at very low levels. These results are in contrast with the conclusions of other reports, which state that RBD-reactive antibodies are not detected in unexposed individuals ^17,20^. However, in these studies the assays were run at higher sample dilutions and therefore low signal may have been missed or misinterpreted as noise. While these RBD-reactive antibody levels are low in the blood, it is possible that they are present in higher concentrations in other sites, such as the mucosa. Also, as antibodies to RBD are associated with virus neutralization both in vitro and in animal models, ^18,33-35^ performing detailed functional analyses of plasma samples from pre-pandemic samples with RBD-reactive antibodies is an important next step. Preliminary experiments from our group indicated that neutralization activity was not present in four subjects (data not shown) but future experiments are needed to more thoroughly address this question.

Individuals over 70 years of age possessed lower levels of both eCoV and cross-reactive SARS-CoV-2 antibodies (Figure 4C). A study comparing pre-existing SARS-CoV-2 reactive T cell immunity in different age groups found that levels were lower with older age ^36^. As individuals over 70 are more likely to present with serious COVID-19 complications ^37-40^ future research investigating connections between age, eCoV and SARS-CoV-2 reactive immunity, and vulnerability to severe COVID-19 is warranted.

Direct comparison of our ELISA protocol with two commercially available serological assays for SARS-CoV-2, Antagen’s LFD test and Abbott’s CMIA IgG assay yielded interesting results. Both the LFD and CMIA performed well in identification of the convalescent subjects via detection of SARS-CoV-2 IgG to RBD (Antagen test) and N (Abbott test), and the BU ELISA detected signal from many pre-pandemic samples which all scored negative in the Antagen and Abbott tests (Table II). However, these commercial tests are specifically designed to detect SARS-CoV-2 infection, unlike the BU ELISA which is measuring all SARS-CoV-2 reactive antibodies; therefore, it is possible the antigens have been modified in the commercial assays to minimize cross-reactive antibody detection, designated as noise in these tests. Interestingly, of the 10 acute subjects that scored negative on the Abbott test, five scored positive for IgM and/or IgG by the LFD test. Also, of the six parameters measured by the BU ELISA, AU values were higher than all pre-pandemic samples for at least one readout for 9/10 subjects. Taken together, these results indicate multi-parameter detection of SARS-CoV-2 reactive antibodies with sensitive tests may improve use of serologic data for diagnostics.

The BU ELISA protocol enables the measurement of low levels of antigen-specific antibodies within high concentration human specimens. Use of this assay could provide new insight into viral transmission and help elucidate the nature of the virus-specific antibody response. Also, this protocol may complement other tests for diagnostics, measurements of COVID-19 vaccine responses, screening of convalescent plasma for clinical use, and perhaps most importantly, to accurately determine a history of exposure to SARS-COV-2.

## Material and Methods

### Participants

#### Pre-pandemics

Samples were collected for unrelated studies prior to December 2019. <u>Acutes</u>: de-identified samples from hospitalized patients at Boston Medical Center with confirmed PCR positivity for SARS-CoV-2. Samples were collected at various timepoints after onset of symptoms. <u>Convalescent:</u> Subjects were recruited by contacting individuals who had been confirmed to have SARS-CoV-2 infection through their exposure at a biomedical conference in March 2020. None were hospitalized. Samples were collected 3-6 months after positive SARS-CoV-2 PCR test. Samples were collected and/or used in this study with proper IRB approval from the Boston University Institutional Review Board.

### The BU ELISA Protocol

Antibodies reactive to all four eCoV2 and SARS-CoV-2 RBD or N were assayed from sera or plasma as described in accompanying SOP (Supplemental Figure 1). Briefly, wells of 96-well plates (Pierce 96-Well Polystyrene Plates; cat#15041, Thermo Fisher Scientific) were coated with 50μl/well of a 2μg/ml solution of each respective protein in sterile PBS (Gibco) or with PBS only for 1 hour at room temperature. Coating solution was removed manually by a swift flick of the plates into a biohazard waste container. Next, 200μl per well of sterile PBS was added with a multichannel pipettor and liquid was removed via swift flick and the plate was banged on absorbent paper towels to remove residual liquid; this washing procedure was performed three times. Next, 200μl of casein blocking buffer (Thermo Fisher Scientific, cat#37528) was added to wells at room temperature for 1 hour. Next, plates were washed three times as previously described. Subject samples and monoclonal SARS-CoV-2 RBD reactive antibodies (IgG, clone CR3022, gift from the Ragon Institute; IgA, clone CR3022, Absolute Antibodies; IgM, clone BIB116, Creative Diagnostics) were diluted in Thermo Fisher casein blocking buffer, and 50μl of each were added to the plates for 1 hour at room temperature, with dilution buffer only added to blank wells. After incubation, samples were removed by a swift flick into a biohazard waste container. The plates were again washed three times with PBS containing 0.05% Tween 20 (PBST) and banged on absorbent paper towels, and immediately anti-human horseradish peroxidase (HRP)-conjugated secondary antibodies for IgG (cat#A18817, Thermo Fisher, 1:2000), IgM (cat#A18841, Thermo Fisher, 1:8000), and IgA (Jackson Immunoresearch, cat#109-035-011, 1:2000) diluted in casein blocking buffer were added to the plates at 50μl per well for 30 minutes at room temperature. Next, plates were washed four times with 0.05% PBST as described, and 50μl per well of 3,3’,5,5’-Tetramethylbenzidine (TMB)-ELISA substrate solution (Thermo Fisher Scientific, cat# 34029) was added and incubation occurred in the dark until a visible color difference between the well with the seventh dilution (1.37ng/ml) of recombinant antibody and the diluent only ‘zero’ well appeared, this time ranged from ∼8-20 minutes. The reaction was stopped by adding 50μl of stop solution for TMB (Thermo Fisher Scientific, cat#N600) and the optical density was measured 450 nm (OD 450nm) on a SpectraMax190 Microplate Reader (Molecular Devices). Seven-point sample dilution curves were run in uncoated wells and paired antigen coated wells (SARS-CoV-2 RBD and NP). An example of a plate map shown in Supplemental Figure 1.

### Antigens

SARS-CoV-2 RBD was a gift from the Schmidt lab at the Ragon Institute and was expressed and purified as previously described ^41^. SARS-CoV-2 N (Cat# 40588-V08B) and S (Cat# 40591-V08H), NL63 N (Cat# 40641-V07E), 229E N (Cat# 40640-V07E), OC43 N (Cat# 40643-V07E) and HKU1 N (Cat# 40641-V07E) was purchased from Sino Biological. Histidine-tagged NL63 and HKU1 RBD sequences were inserted into plasmid vector VRc (gift from the Schmidt lab at the Ragon Institute) and was expressed in 293 Freestyle cells (293F, ThermoFisher) and purified on Ni-NTA resin as previously described ^42^ (ref).

### Determination of Arbitrary Units

Data were analyzed using GraphPad Prism 8. Arbitrary units (AU) on a ng/ml scale were calculated from the optical density (OD) values according to standard curves generated by known amounts of monoclonal anti-SARS-CoV-2 RBD IgG, IgM, or IgA. The OD values of blank (diluent only) wells with the same coat and secondary detection antibody were averaged and subtracted from the OD values of each respective sample well and then the ODs were logarithmically transformed. Next, a non-linear regression of the sigmoidal standard curve was used to extrapolate a “concentration” for the patient samples, which was then inverse log transformed and multiplied by the respective dilution factor. AU values for each sample were chosen from the linear portion of the dilution curve for the antigen coated wells, and the paired buffer only coat value was subtracted to determine the net AU amount.

### Determination of the presence versus absence of SARS-CoV-2 reactive antibodies in samples and of Arbitrary Unit Values

First, the average ODs of corresponding ‘blank’ wells (sample diluent only in buffer only coated or antigen coated) on a given plate was subtracted from all wells with samples. ODs for blank wells was consistently ∼0.05 regardless of coat. <u>Metric 1</u>: Signal was considered positive from a given subjects if (1) the OD values from the antigen coated wells was a minimum of 2.5x higher than that of the paired buffer coated well for at least two sample dilutions and (2) one antigen-coated well OD value was over 0.1, after the average OD values of the respective blank wells were subtracted.

### LFD tests

Antagen’s DISCOVID IgM IgG LFD test was used to detect SARS-CoV-2 RBD specific IgM and IgG antibodies following manufacturer instructions. Briefly, 20μl of plasma or serum was added to the indicated sample port, immediately followed by provided diluent, and incubated at room temperature before reading at 45 minutes. The results were scored as positive or negative for IgM and IgG by two independent readers blinded to donor sample status.

### Abbot Serology Test

The SARS-CoV-2 IgG assay is a chemiluminescent microparticle immunoassay (CMIA) used for the detection of SARS-CoV-2 nucleocapsid protein-specific IgG in human samples. The assays were performed according to manufacturer’s protocol.

### Automated Washer

Plates were washed with Molecular Devices SkanWasher 400 microplate washer with three rounds of aspiration and wash with a final aspiration step for each run. This protocol was run twice after the coating, blocking, and sample incubation steps and three times after the addition of the secondary detection antibody step in the experiment shown in Figure 1A. Plates were rotated 180° between each run. Residual wash buffer was left in the plates (plates were not blotted post-wash) to mimic a fully automated system.

## Data Availability

The raw data supporting the conclusions of this manuscript will be made available by the authors, without undue reservation, to any qualified researcher.

## Author Contributions

JES-C conceived of the experimental plan, RY and JES-C designed the approach, AJC, RY, JES-C designed experiments; RC, ES, NL, KQ, PU, NB, GM, WG, IR, MS, JES-C contributed to study subject specimen collection/use, provided reagents, and/or funding; RY, RP, DS, EC, JES-C, AO, LB, FK, JB, AB, YK performed experiments; RY, DS, EC, JB, SG, AB, GM, YK, and JES-C analyzed data; and RY and JES-C wrote the manuscript.

## Acknowledgements

We thank Dr. Aaron Schmidt/Dr. Jared Feldman and Dr. Galit Alter from the Ragon Institute of MGH, MIT, and Harvard for the gifts of the recombinant SARS-CoV-2 RBD antigen and monoclonal SARS-CoV-2 RBD IgG antibody, respectively. We thank Dr. Wenda Gao for the gift of the Antigen rapid COVID antibody (LFD) tests. We also thank Dr. Andrew Lodge at ThermoFisher Scientific for his advice regarding initial reagent selection and protocol steps. The manuscript has been released as a pre-print at MedRxIV, (Yuen et al)^43^

## Title: BU ELISA protocol for the detection of IgG, IgM, and IgA isotypes reactive with SARS-CoV-2 RBD

**Date:** December 16^th^, 2020

### Prepared by

Jennifer Cappione and Rachel Yuen

### Purpose

The purpose of this SOP is to provide a protocol for detecting the levels of IgG, IgM, and IgA SARS-CoV-2 RBD-specific antibodies in human serum and plasma samples.

### Requirements

Liquid biological samples (serum or plasma) from human subjects

### Equipment

1. Biological Safety Cabinet – Class II or higher
2. Pipette aid
3. Single and Multi-channel pipettors: 1-1000ul
4. ELISA plate reader
5. Waste containers (i.e. bleach buckets, biohazard buckets)
6. Vortex

### Consumables

1. Serological Pipettes (5ml-25ml)
2. Pipette tips, sterile (10-1000ul)
3. ELISA plates (Pierce 96-Well Polystyrene Plates, corner notch; ThermoFisher, cat#15041 or Thermo Scientific(tm) Clear Flat-Bottom Immuno Nonsterile 96-Well Plates (Immulon 2HB); ThermoFisher, cat #3455)
4. Plate sealers (ThermoFisher, cat#3501)
5. Polypropylene tubes (1.2ml-15ml)
6. Reagent reservoirs
7. Absorbent paper towels

### PPE

1. Lab coats that can fully button
2. Disposable nitrile gloves
3. Disposable cuffs

### Reagents and Reagent Preparation

Sterile reagents and sterile technique with proper BSL2 practices are required. Reagents should be stored as recommended by the manufacturer and only used until expiration date. Discard reagents that appear to have contamination.

1. Sterile 1x PBS, without calcium and magnesium (Gibco, cat# 14190144) or similar
2. Tween 20
3. Recombinant SARS-CoV-2 Receptor Binding Domain (RBD), recombinant (gift from Schmidt lab at Ragon Institute)
4. Casein blocking buffer in PBS (ThermoFisher, cat# 37528)
5. Anti-SARS-CoV-2 Monoclonal Antibodies: CR3022 IgG (gift from Ragon/Harvard), CR3022 IgA (Absolute Antibody, cat# Ab01680-16.0), BIB116 IgM (Creative Diagnostics, cat# CABT-CS044)
6. Wash Buffer 1
  a. Sterile 1x PBS, no calcium and no magnesium
7. Wash Buffer 2
  a. Sterile 1x PBS, no calcium and no magnesium
  b. 0.05% Tween 20
8. HRP-conjugated anti-human antibodies (IgG, ThermoFisher, cat# A18817; IgM, ThermoFisher, cat# A18841; IgA, Jackson ImmunoResearch, cat# 109-035-011)
9. TMB-ELISA substrate solution (ThermoFisher, cat# 34029)
10. Sulfuric acid stop solution (ThermoFisher, cat#N600)
11. Disinfectant (i.e. 70% v/v ethanol disinfectant – spray bottle, 10% v/v bleach or 10% v/v Wescodyne – bucket or beaker)

### Procedure

All work should be performed inside biological safety cabinets (BSC), level 2 or higher, at room temperature (15-30°C).

Spray down all surfaces, racks, and reagent bottles with 70% v/v ethanol prior to entering and using BSC.

*See Figure 1 for example of experimental plate layout*.

Step 1: Coating plates

1. Dilute stock RBD to 2μg/ml in sterile 1x PBS in polypropylene tubes or directly in reagent reservoirs.
  a. Vortex stock RBD for four seconds at max speed prior to removing aliquot for dilution. Next, mix coating solution with RBD thoroughly by pipetting 8 times with 10ml pipette or by vortexing if using polypropylene tubes.
2. Plate 50μl per well of prepared RBD or PBS as coating control with sterile tips, using caution to not touch the bottom of the wells.
3. Cover tightly with a plate sealer and gently tap plates on all sides, while rotating, to ensure complete coating of the wells.
4. Incubate for 1 hour at room temperature.

Washing Step - (3 times with Wash Buffer 1 unless stated otherwise)

#### Washing Step Details

Perform washing step one plate at a time using multichannel pipettors.

1. Invert plate and flick out coating solution/blocking buffer/samples into waste container (shallow rimmed Tupperware, for example) lined with dry paper towels to prevent splashing of liquid.
2. Immediately add 200μl of washing solution to each well, again careful to not touch the bottom of the wells with the pipette tips.
  a. When wash buffer is added to all wells necessary, lift plate and gently tap 3 times to mix a bit, allow wash buffer to sit in plate for ∼30 seconds, then repeat gentle tapping, and then flick out wash buffer into paper-towel lined waste container and bang plates on <u>dry</u> paper towels 3 times, rotate 180 degrees, bang 3 more times, ensure liquid is removed but do not allow them to sit dry.
3. Repeat step 2 for a total of 3 times, unless stated otherwise.

Step 2: Blocking plates

1. Plate 200μl of cold casein blocking buffer per well.
2. Seal plate with a plate sealer and allow to incubate for 30 minutes – 1.5 hours at room temperature.

Washing Step-(as described above, 3 times with Wash Buffer 1)

Plates can sit in Wash Buffer 1 until ready to add samples and must not be left dry.

Step 3: Add Samples

Samples can be either serum or plasma. If stored at ≤-20°C, allow to thaw completely at 4°C or on ice prior to use. Recommended to not use samples that have undergone more than one freeze-thaw cycle.

1. Prepare samples and standards prior to adding to the plate in separate polypropylene tubes; cluster tubes or microcentrifuge tubes, for example.
  a. Dilute samples in casein blocking buffer to 1:5, 1:10, 1:25, 1:125, 1:625, 1:3,125, 1:30,125 (dilutions may vary per experiment). Prepare standard curves using a 3-fold serial dilutions, starting at 1000ng/mL, in casein blocking buffer.
  b. Mix thoroughly by pipetting or vortexing between each dilution and using new tips every time moving liquid from one dilution stock to another. * Recommended to transfer prepared samples and standards to cluster tubes for ease of transfer to plates.
2. Add 50μl of prepared standard and sample dilution per well, as well as 50μl of casein blocking buffer per well as a negative control for each sample (see example plate map below). * Add samples quickly, gently pipette to mix each sample briefly prior to transfer to plates, using caution to not touch the bottom of the wells and change tips between samples.
3. After all samples are added to a plate, seal with a plate sealer and check for complete coating of the wells; if necessary, gently tap plates on all sides, while rotating.
4. Allow to incubate for 1 hour at room temperature.

Washing Step (as described above, 3 times with Wash Buffer 2)

Plates can sit in Wash Buffer 2 until ready to add samples and must not be left dry.

Step 4: Add HRP-conjugated antibodies

1. Prepare HRP-conjugated antibodies using casein blocking buffer as diluent in polypropylene tubes. Mix stocks and diluted solutions thoroughly by pipetting or vortexing.
  a. IgM – 1:8,000
  b. IgG – 1:2,000
  c. IgA – 1:2,000
2. Add 50μL per well of prepared antibodies to corresponding wells.
3. Seal with a <u>new</u> plate sealer and check for complete coating of the wells; if necessary, gently tap plates on all sides, while rotating.
4. Allow to incubate for 30 minutes at room temperature.

Washing Step (as described above, 4 times with Wash Buffer 2)

Plates can sit in Wash Buffer 2 until ready to add samples and must not be left dry.

Step 5: Development of Plates and Addition of Stop Solution

1. Add 50μl of TMB-ELISA substrate solution to each well. * TMB-ELISA substrate solution should be equilibrated to room temperature prior to use. * Check for complete coating of the wells; if necessary, gently tap plates on all sides, while rotating.
2. Allow to incubate for decided number of minutes in the dark, unsealed (ranging 8-20 minutes or when visible color difference is seen between most diluted standard (dilution 7) and casein blocking buffer only well, but multiple reads at 652nm can be taken before stopping assay to ensure best signal:noise is achieved).
3. Stop reaction by adding 50μl of sulfuric acid stop solution to each well.

Step 6: Read Plates on Microplate reader

(1) Measure absorbance at 450nm.

**Figure 1.**
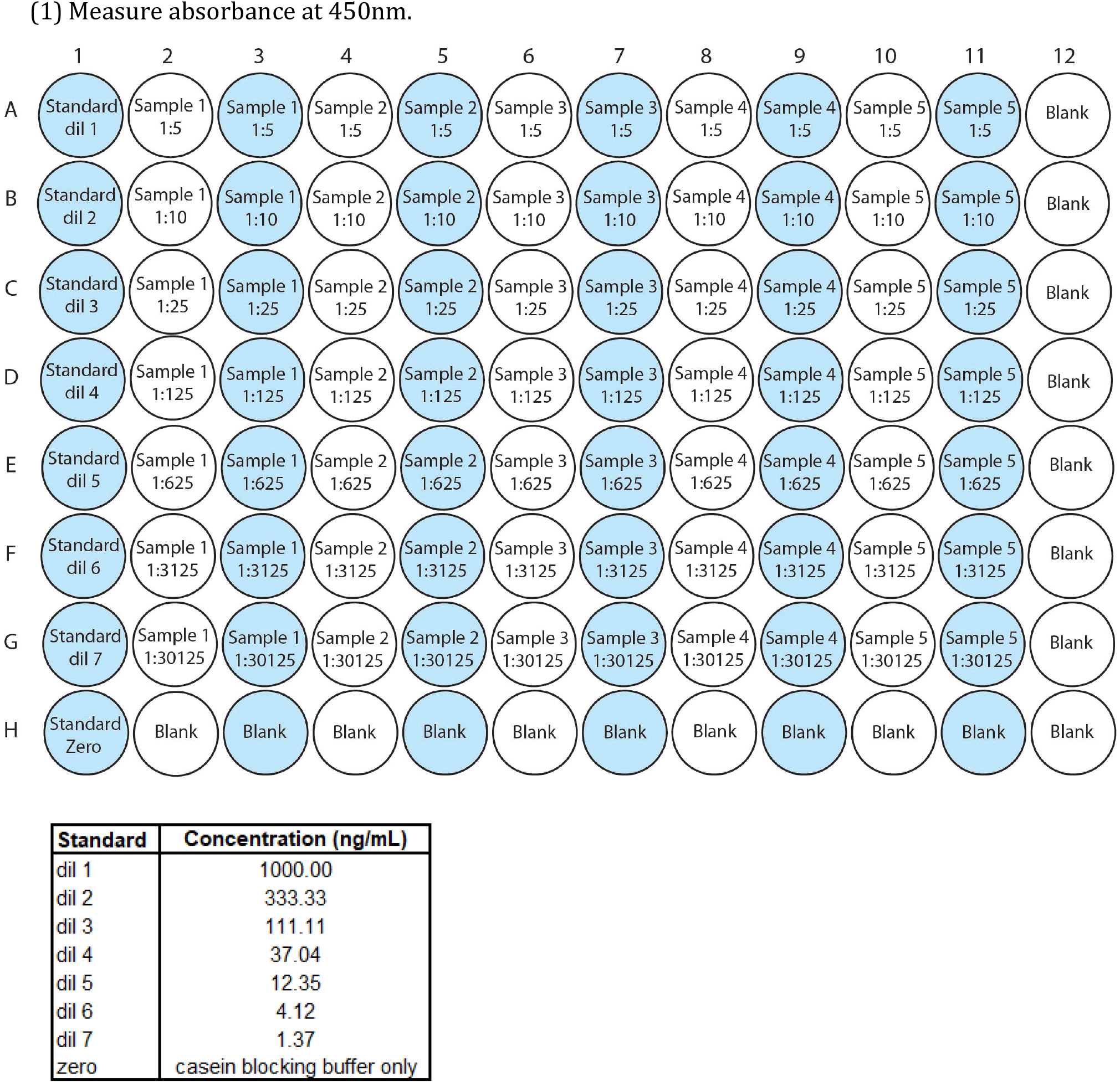
Example experimental plate layout.

**Supplementary Figure 2.**
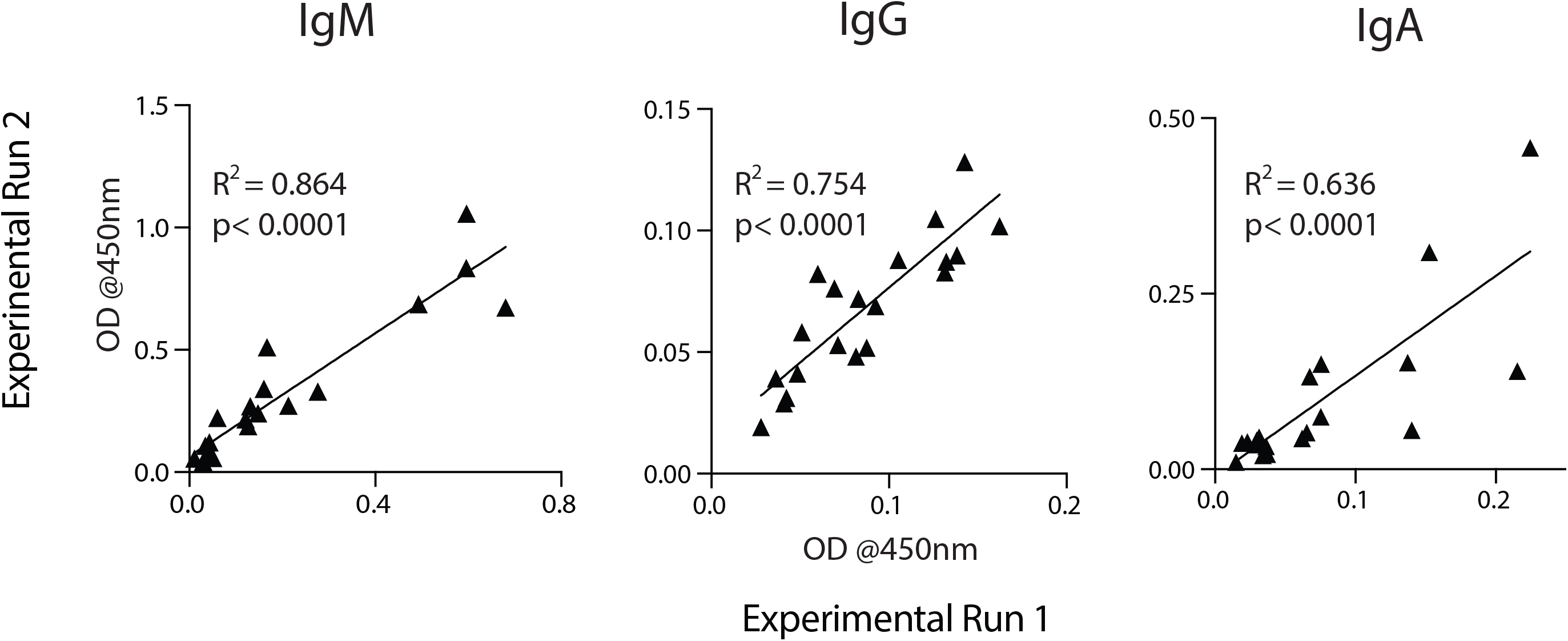
The OD values from subject samples from buffer only coated wells are highly consistent between experimental runs. OD values of a 1:5 sample dilution from 34 representative subjects for IgM, IgG and IgA from two different experimental runs with the BU ELISA protocol are shown. Pearson’s rank test and linear regression analysis was performed, and R-squared and p values are shown.

**Supplementary Figure 3.**
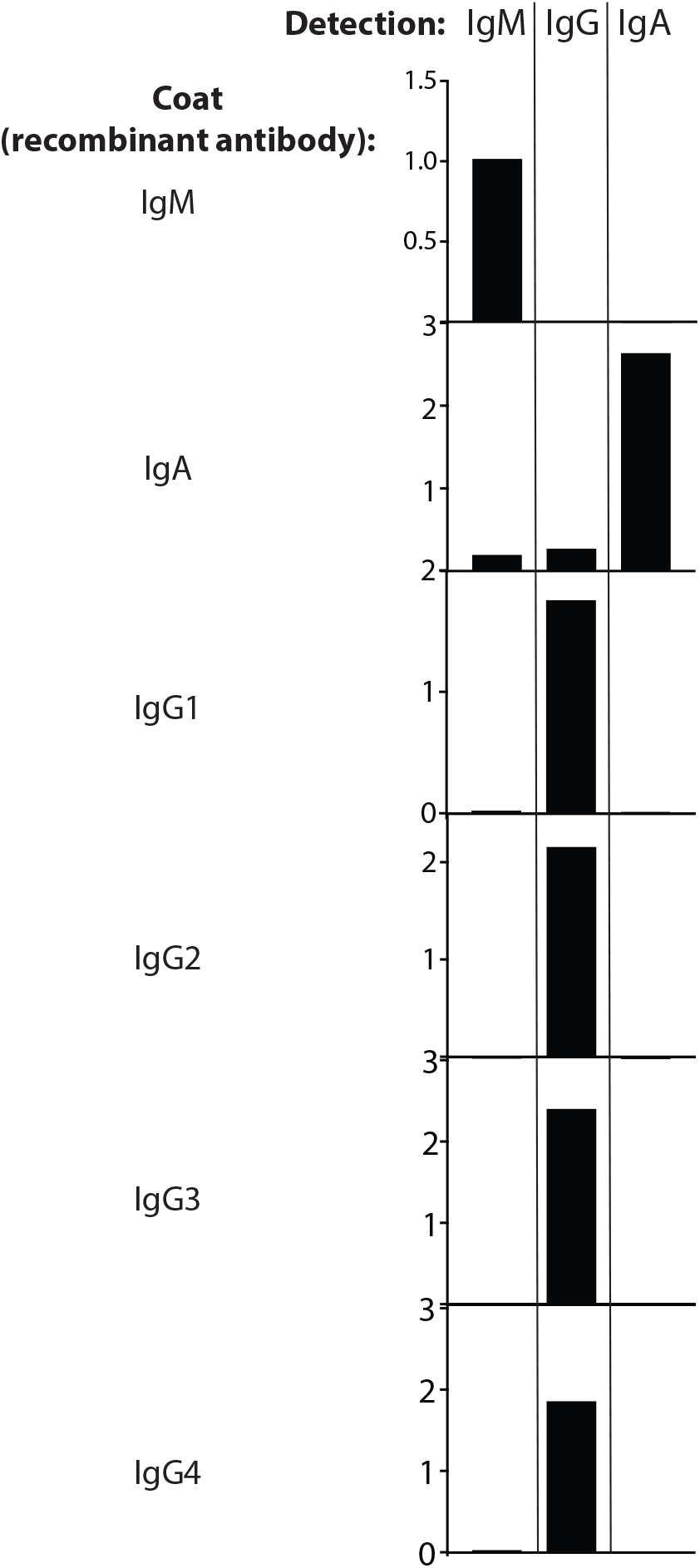
Test of specificity of detection antibodies. Plates were coated with monoclonal recombinant antibodies and probed with corresponding detection antibodies. Experiment was performed once.

**Supplementary Figure 4.**
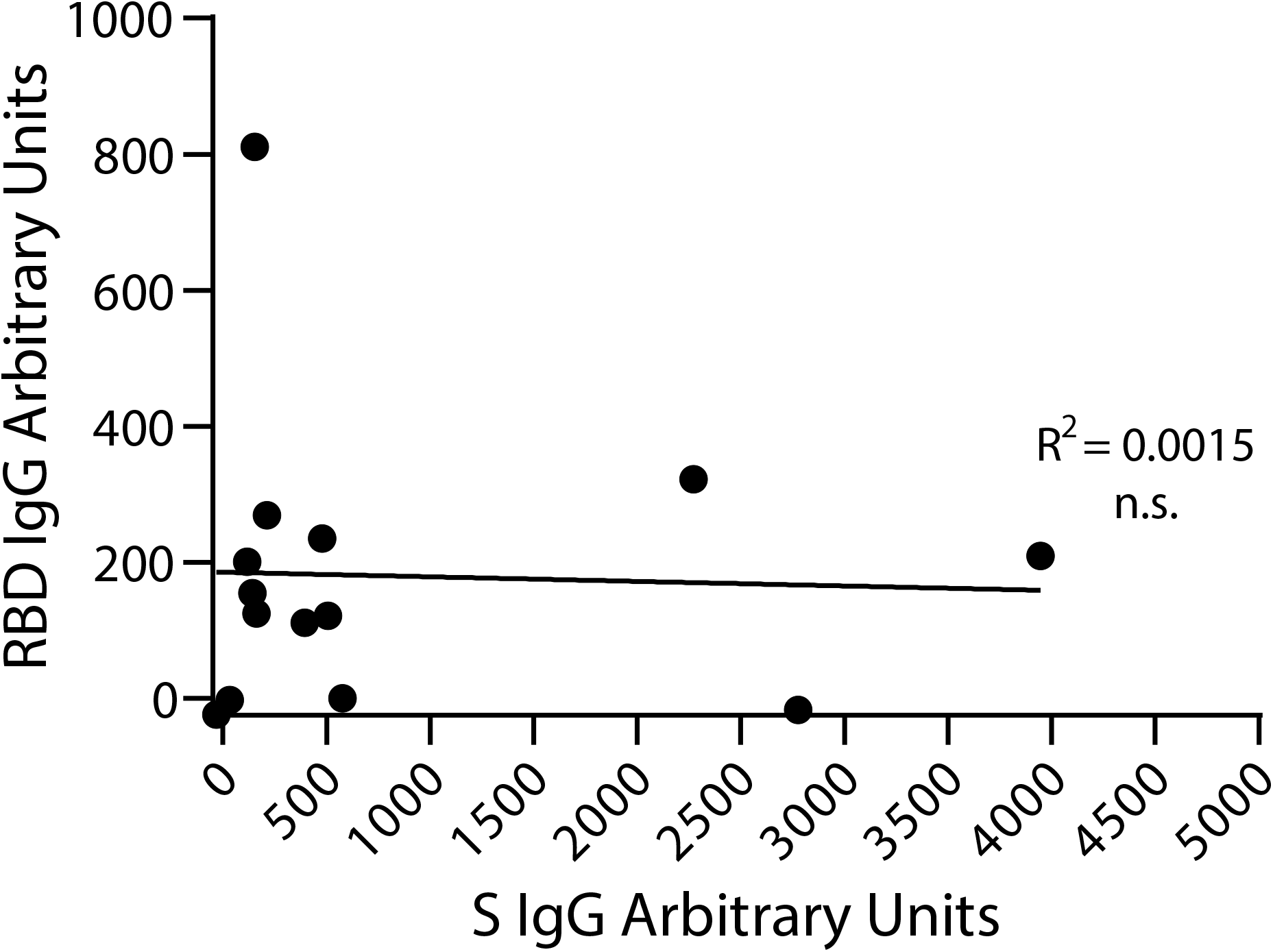
Reactivity of pre-pandemic samples to SARS-CoV-2 S and RBD. Correlation between IgG reactive to SARS-CoV-2 S and RBD in pre-pandemic samples (n = 14). Pearson’s rank test and linear regression analysis was performed, and R-squared and p value is shown.

